# Development of Diagnostic Real-Time Reverse Transcription-PCR Assays for SARS-CoV-2 using Polyvinyl Alcohol (PVA) Sponge as Saliva Collection Tool

**DOI:** 10.1101/2025.11.24.25340856

**Authors:** Kenji Kinoshita, Madoka Kisoi, Kazumasa Shinozuka, Tadahiro Sasaki, Tatsuo Shioda, Atsushi Ichikawa

**Affiliations:** PCR Center, Mukogawa Women’s University, Kyuban-cho, Koshien, Nishinomiya, 663-8179, Japan; Department of Viral Infections, Research Institute for Microbial Diseases, Osaka University, 3-1 Yamadaoka, Suita, Osaka 565-0871, Japan; Bio Education Laboratory, 1-21-33 Higashinakajima, Higashiyodogawa, Osaka, 533-0033, Japan

**Keywords:** SARS-CoV-2, Saliva, Direct RT-qPCR, PVA sponge, Sample pooling method

## Abstract

Given the pandemic of COVID-19, the diagnostic challenges using Reverse Transcription-quantitative Polymerase Chain Reaction (RT-qPCR) assays for SARS-CoV-2 are mainly concentrated in the operation, which is to timely process multiple samples and to judge false-positive/false-negative accurately. Here we suggest a unique real-time RT-PCR protocol using a polyvinyl alcohol (PVA) sponge for high-throughput screening against COVID-19. By collecting saliva using a PVA sponge, we have solved operational issues allowing for skipping of conventional sample preparation and purification procedures. While standard sample pooling methods can be time consuming, costly, and the expected positive rate of the sample can be low, our method allows for simplification of the procedures for pool testing. Moreover, the decrease in sensitivity due to dilution when the solution is added can be prevented when testing samples using pieces of dried PVA sponges. Ribosomal protein L-13A (*RPL13A*), a human-derived housekeeping gene of internal control, are included in the primers and probes of the assay kit for simultaneous detection. Therefore, the accuracy for avoiding false-positive or false-negative RT-qPCR assay results can be improved. In summary, we have developed a novel RT-qPCR testing procedure by uniquely using PVA sponge without purification, which can be utilized for the detection of SARS-CoV-2.

**Graphical Index:** 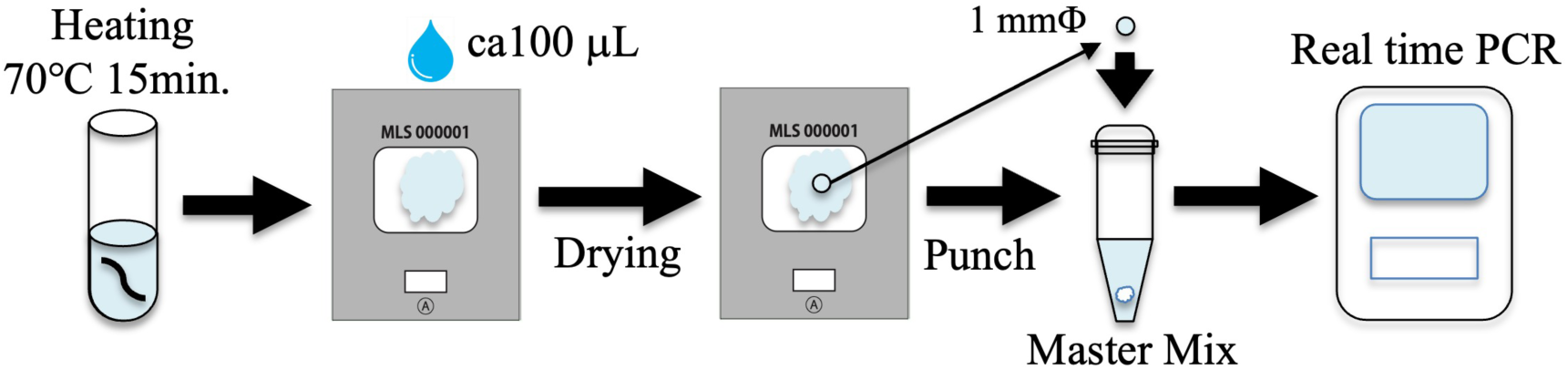

## Introduction

<u>S</u>evere <u>A</u>cute <u>R</u>espiratory <u>S</u>yndrome-<u>Co</u>rona<u>v</u>irus-2 (SARS-CoV-2) has quickly spread globally^1^. The <u>Co</u>rona<u>vi</u>rus <u>D</u>isease 20<u>19</u> (COVID-19) pandemic has created a major demand for accurate PCR tests that detect viral infection caused by SARS-CoV-2. <u>R</u>everse <u>T</u>ranscription-<u>q</u>uantitative <u>P</u>olymerase <u>C</u>hain <u>R</u>eaction (RT-qPCR) has been utilized to confirm the viral load of the SARS-CoV-2. In order to handle clinical samples and perform SARS-CoV-2 and other excluded experiments, inactivation methods are generally required to work under safe conditions. In addition, virus inactivation allows the movement of materials from biosafety level 3 (BSL-3) to the BSL-2 laboratory, promoting experimental performance and performing experiments. Moreover, virus-containing biological specimens collected from subjects contain components that negatively affect detection of viral RNA by degrading RNA and inhibiting RT-PCR. Several methods for virus inactivation are available, and the choice of which approach to use is related to their compatibility with downstream applications. Heat inactivation has been used for several viruses^2,3^ and is a common method employed for antigen preservation of viral and bacterial pathogens. Moreover, in direct RT-qPCR tests, treatment of non-ionic detergents, such as Tween20, to saliva has been reported to increase the sensitivity of SARS-CoV-2 derived RNA^4–6^.

Commonly used tests to detect viral RNA involve removal of extraneous components from a biological specimen by RNA purification. The application of qPCR for the relative quantification of an RNA of interest which is preceded by first step is the isolation and purification of total RNA from the specimens. The second step follows the use of a single-reaction kit combining the RT and qPCR reactions. Specifically, the National Institute of Infectious Diseases (NIID), Japan protocols require the use of RNA purification kits, which not only results in a significant cost increase but leads to a major supply shortage of such kits. Recently, several RT-qPCR test kits in COVID-19 detection have been developed that eliminate the viral RNA purification step, which is a minimally processed virus-containing specimen. Direct RT-qPCR has provided several advantages for streamlining diagnosis because it is faster and easier. The most direct RT-qPCR tests use saliva because it is a more sensitive detection tool for SARS-CoV-2, and it is simpler and safer to collect^7–11^ clinical specimens containing viral particles are inactivated by heating or by dissolving them directly in a buffer containing a detergent. The inactivated sample will be used for the next direct RT-qPCR diagnostic reaction. However, direct lysis in a detergent containing buffer promotes the degradation of SARS-CoV-2 viral RNA by RNases. Therefore, it is necessary to process the sample at low temperature and proceed directly to the RT-qPCR reaction as soon as possible.

Theoretically, if efficient and accurate diagnostic assays were available to identify infections, rapid isolation of infected individuals before the viral load would become high and prevent further infection chains. To achieve such an ideal scenario in a large population, there are several operational issues to overcome. One of the issues is that the processing procedure using RT-qPCR can be labor intensive because of the need to collect high purity samples, as well as the need of sophisticated facilities with trained experts^12, 13^.

To reduce the labor intensive required for operation, there is a need for improvements to simple and quick operation methods. From that point of view, we have been developing devices and genetic testing methods that enable genetic testing from various samples such as blood, saliva, hair roots, and food without conventional genomic DNA/RNA extraction and purification^14–17^.

Our goal was to further simplify detection of SARS-CoV-2 viral RNA by direct RT-qPCR and to enhance its sensitivity. We focused on the initial processing of clinical specimens because this step is time-consuming and critically affects the sensitivity of subsequent RNA detection assays. Here, we describe a novel direct RT-qPCR based method for the detection of SARS-CoV-2 using a highly biocompatible **<u>P</u>**oly **<u>V</u>**inyl <u>A</u>lcohol (PVA) sponge compared to other direct methods. This method simplifies the conventional method, while maintaining its sensitivity and specificity. As a result, with our ultimate goal of providing convenient, scalable, and cost-effective direct RT-qPCR for >1,000 individuals per day at a PCR center, we here report the discovery of a sensitive saliva-based detection method for SARS-CoV-2. This SARS-CoV-2 testing process and workflow is convenient, simple, rapid, and inexpensive, and can be readily adopted by any testing facility currently using real-time PCR equipment.

## Experimental

### PVA Sponge

The PVA sponge has a porous structure made from water soluble PVA acetalized with an acid catalyst. The finished product becomes Polyvinyl formal. During the acetalizing process, the starch is added as pore forming agent. After a water-soluble porous structure is made, the starch is extracted by water. The PVA sponge referred to as polyvinyl alcohol formalized sponge, is a high qualities plastic sponge that has many of the same properties and qualities of a natural sea sponge. The PVA sponge used in this study is highly biocompatible and can be used as a material to promote the real-time PCR reaction. Saliva was dropped on the PVA sponge, and dried overnight at room temperature. A small piece of the PVA sponge was added to the master mixture to proceed with direct RT-qPCR. By using a PVA sponge, we have overcome operational challenges allowing the process to be simplified from conventional sample collection to a purification procedure.

### Prototype Sampling Kit UBS Card

The UBS Card (Universal Bio-Sampling Co. Ltd., Japan) was designed with a structure that took into account processes such as saliva sampling, drying, sample punching, mailing, and storage as shown in Fig. 1. A plastic plate (polyethylene; PE) for a punch stand was attached to the bottom of the PVA sponge (1.0 mmT). The UBS Card covered the upper part of the PVA sponge and has a space of ca 5 mm on both sides to prevent the risk of contamination and to promote drying before and after application of the saliva sample. The sampling kit UBS Card is designed so that the PVA sponge part can be cut off in consideration of the automation of punching work.

**Fig. 1.**
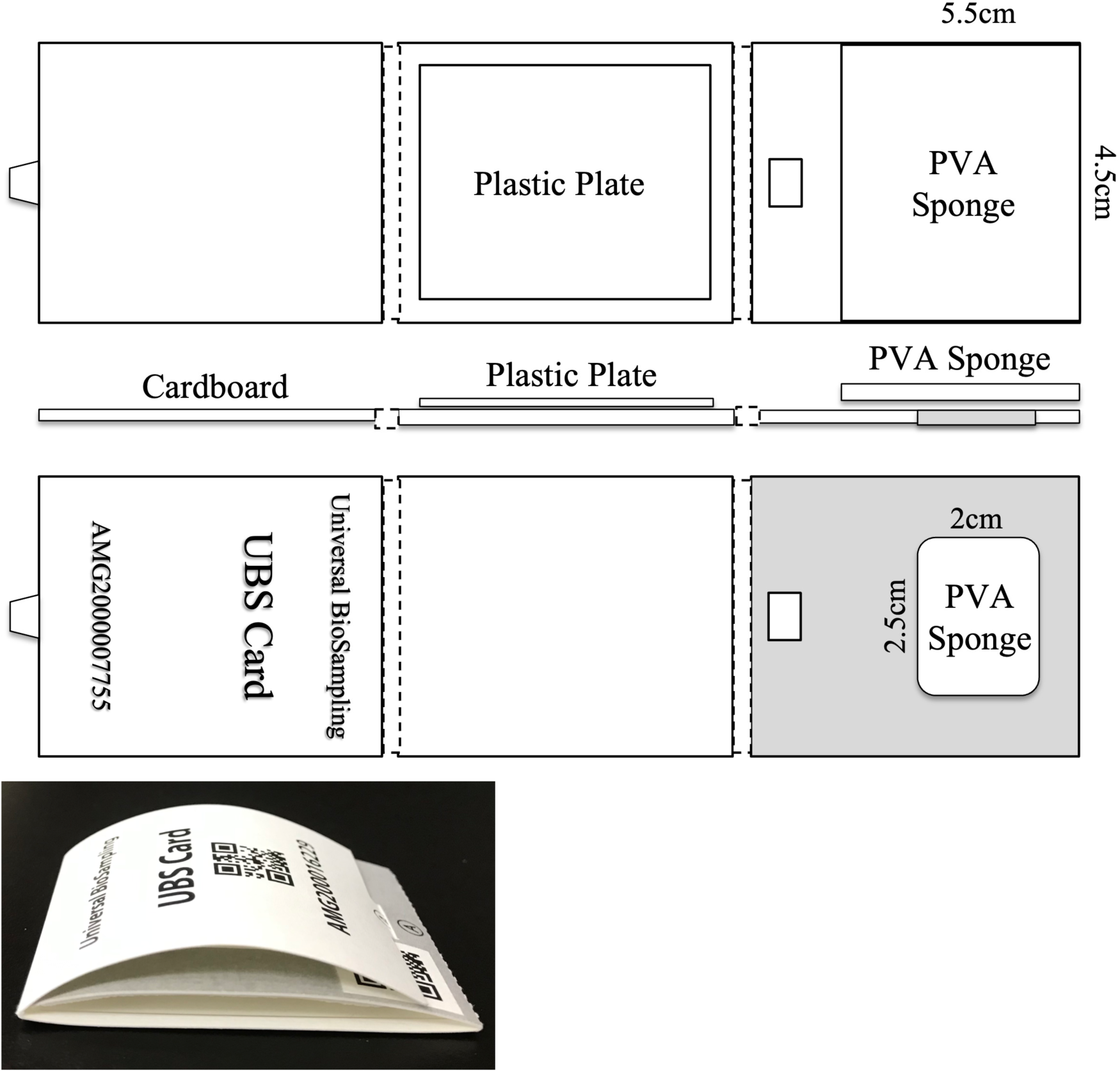
Design of the Homemade Sampling Kit “UBS Card”

### Preparation for Standard SARS-CoV-2 and Quantification

Vero E6 cells were maintained in Eagle’s minimum essential medium (MEM) containing 10% fetal bovine serum (FBS), and antibiotics (100 units/ml penicillin and 100 μg/ml streptomycin) at 37°C with 5% CO_2_. SARS-CoV-2 JPN/TY/WK-521 was obtained from the <u>N</u>ational <u>I</u>nstitute of <u>I</u>nfectious <u>D</u>iseases (NIID), Japan and was cultured in Vero E6 cells with MEM containing 2% FBS and antibiotics at 37°C with 5% CO_2_ and was harvested 48 h post inoculation. The supernatant of the virus culture was centrifuged at 3000 rpm for 10 min at 4°C, and aliquots were then stored at −80°C. For confirmation of viral heat inactivation, 0.5 ml of the viral stock was pipetted into 2 ml safe-lock micro centrifuge tubes, which were incubated at 70°C for 15 min. According to previous paper^18^ the number (titer) of infectious SARS-CoV-2 in samples were measured using the **<u>M</u>**edian **<u>T</u>**issue **<u>C</u>**ulture **<u>I</u>**nfectious **<u>D</u>**ose 50 (TCID_50_) assay, which is a rapid and simple focus-forming assay using the peroxidase-antiperoxidase technique. Using TCID_50_ assay, we can measure the number of infectious virus particles up to the number of 2.0×10^1^ virus particles/ml. The virus number values were compared with the cycle threshold (*Ct*) values of real-time RT-qPCR. All the experiments using SARS-CoV-2 were carried out in the BSL-3 laboratories of the Research Institute for Microbial Diseases of Osaka University.

The day before infection, 2.0 x 10^4^ cells per well were seeded in a 96-well tissue culture plate using MEM containing 10% FBS and antibiotics and incubated overnight at 37°C 5% CO_2_. The cell supernatant was removed, and 50μL of MEM containing 2% FBS and antibiotics was added to the plate. Serial 10-fold dilutions of the samples for virus titration were prepared in MEM containing 2% FBS and antibiotics, and each dilution was inoculated to the cells in six replicates. The plates were incubated for 72 h at 37°C, 5% CO_2_. Then, the cells were fixed with 7% formaldehyde PBS and stained with 1% methylene blue solution. Virus titer in a TCID_50_ assay was calculated according to the Reed and Muench method based on six replicated for the dilution as previously described^19^.

### One-step Direct RT-qPCR for SARS-CoV-2 Viral RNA

Unique real-time RT-qPCR tests were performed using the QuantStudio™ 12K flex real-time PCR system TaqMan^®^ Genotyper^TM^ software (Thermo Fisher Scientific Co., Ltd.). The assays were performed with SARS-CoV-2 RT-qPCR Detection Kit Ver.2 (FUJIFILM Wako Pure Chemical Co., Ltd., Japan) according to the manufacturer’s instructions. The RT-qPCR master mixture in a 10 μL reaction were as follows: 0.125 μL of Hot Start Reverse Transcription DNA Polymerase (TTx DNA Polymerase), and 4 μL of Reaction buffer including dNTPs, and 0.5 μL of 50 mM manganese (II) acetate, and 0.5 μL each of Primers and Probes No. 1 and No. 2, and 0.2 μL of ROX ×500 reference dye, and 4.174 μL of distilled water (DW). A small piece of the PVA sponge (1.0 mm<!) which was cut-off by biopsy punch (BPP-10F, Kai Industries Co., Ltd. (Tokyo, Japan) or 1 μL of the liquid specimen was added directly into the PCR tubes. All of the direct RT-qPCR preparations were performed at 4°C. The cycling conditions were 90°C for 5 min for initial denaturation and enzyme activation, 60°C for 10 min for reverse transcription, 95°C for 1 min for the second denaturation, followed by 50 cycles each of 95°C for 10 sec and 65°C for 20 sec, respectively.

### Linear Regression Analyses of the SARS-CoV-2 Standard Solutions

Finally, to experimentally validate the effectiveness of the thermal inactivation procedure, we propagated SARS-CoV-2 in vitro, and subjected active harvested viral particles to heating at 70°C for 15 min. The confirmation of inactivation for SARS-CoV-2 was performed plaque assays on Vero E6 cells.

To assess the quantitative ability of the SARS-CoV-2 RT-qPCR Detection Kit Ver.2, the linearity of the SARS-CoV-2 N1 and N2 gene analyses was confirmed using the SARS-CoV-2 standards (STDs). Standard SARS-CoV-2 viral RNA solutions as positive controls were prepared as follows: Inactivated SARS-CoV-2 (2.7×10^7^ copies/μL) in a Dulbecco’s Modified Eagle Medium was serially diluted with DW to make standard SARS-CoV-2 viral RNA from 2.7×10^5^ copies/μL to 2.7×10^2^ copies/μL. Furthermore, in order to confirm the linearity in saliva, we designed three types of analytical conditions using saliva as follows: The saliva (Normal, Pooled Donors, 991-05-P) was purchased from Lee BioSolutions, Inc. (MO, USA). These sample preparations were also performed at 4°C. The condition was for the purpose of analyzing the RNA STDs in the saliva heat-treated at 70°C for 15 min with the same concentration as above.

1 μL of SARS-CoV-2 viral RNA STDs solution was added directly into the PCR tube at 4°C. Each tube was filled with 10 μL of RT-qPCR master mixture, and then the direct RT-qPCR analyses were subsequently performed using a QuantStudio™ 12K Flex Real-Time PCR System (Thermo Fisher Scientific Co., Ltd.).

The 100 μL of each standard sample prepared in the previous section was added dropwise to the PVA sponge on each UBS card and dried overnight at room temperature in a Class II biosafety cabinet. In order to evaluate the quantitative ability of the SARS-CoV-2 RT-qPCR detection kit using the SARS-CoV-2 STDs PVA sponges adjusted above, the linearity of the previously prepared SARS-CoV-2 STDs solution was confirmed by the following method as shown in Fig. 2. These SARS-CoV-2 STDs solutions were also kept at room temperature until direct RT-qPCR. After 24 h, 1 μL of SARS-CoV-2 viral RNA STDs solution or a small piece (1.0 mmΦ) of the sample was dropped directly into the PCR tube at 4°C. Each tube was filled with 10 μL of RT-qPCR master mixture, and then the direct RT-qPCR analyses were subsequently performed using a QuantStudio™ 12K Flex Real-Time PCR System (Thermo Fisher Scientific Co., Ltd.).

**Fig. 2.**
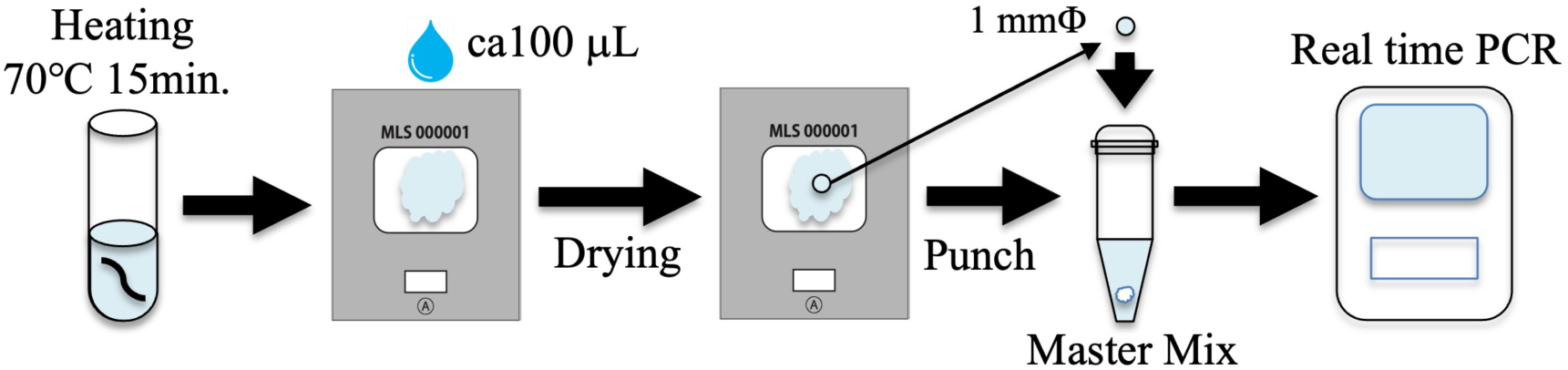
Schematic Overview of SARS-CoV-2 Direct RT-qPCR Testing Procedure. In order to simulate the inactivation of the virus, it was treated by heating. Following this, as a sample preparation, inactivated saliva was dropped on the PVA sponge and then dried overnight at room temperature. A small piece of the PVA sponge was transferred to the PCR tube with the RT-qPCR master mix. For comparison with the PVA sponge method, the virtually inactivated liquid samples were also used for the direct RT-qPCR diagnostic reaction.

Each experiment was performed in triplicate. To confirm the test results of the SARS-CoV-2 genes N1 and N2 analyses, calibration curves were graphically presented from the *Ct* values using the QuantStudio^TM^ 12K Flex Real-Time PCR System TaqMan® Genotyper™ Software (Thermo Fisher Scientific Co., Ltd.).

### General Saliva Sampling and Preparation Using PVA Sponge

The unique direct RT-qPCR based method has been performed at Mukogawa Women’s University as follows (Fig. 5). First, the subject receives a sampling kit, which includes the Trust Catcher (Trust Medical Co. Ltd. Japan) and the Eppendorf tube. The Trust Catcher is maintained in the subject’s mouth for a few minutes until the sponge is sufficiently soaked with saliva. The saliva is then transferred to the Eppendorf tube after the sponge rod is pushed into the container. The saliva in the Eppendorf tube is heat-treated in a heat block at 70 °C for 15 min. The 100 µL of the inactivated saliva samples were dropped on the PVA sponge and then dried in the class II biological safety cabinet at room temperature. Small pieces of the specimens (1.5 mm<!) were punched out from the PVA sponge using the Wallac DBS Puncher (PerkinElmer Co., Ltd., USA), then automatically dropped directly into the PCR tubes. Each tube was filled with RT-qPCR master mix solutions, which were adjusted according to the instruction

### Unique Direct RT-qPCR for Sample Pooling Method

The RT-qPCR master mixture in a 40 μL reaction was adjusted to 4 times the conditions described before in the one-step direct RT-qPCR, and the cycling conditions were the same as those used in the same as before. The PVA sponge specimens (1.0 mm<!), in which the 100 μL of the standard SARS-CoV-2 DW solution 2.7×10^3^ copies/μL was added dropwise to the PVA sponge on the UBS card, dried overnight at room temperature, and then was added directly into the PCR tubes.

## Results and Discussion

### General Diagnosis for SARS-CoV-2 by RT-qPCR

Routinely, quantitative PCR is preceded by the isolation and purification of total RNA from the specimen, and by reaction resulting in complementary DNA (cDNA) from the template RNA which is then utilized for the qPCR reaction. However, the nucleic acid purification is not only laborious and time-consuming, but the additional steps requiring manual handling can be also led to experimental contamination. In the case of clinical sampling and diagnostics, the use of a single reaction kit combining the RT and qPCR reactions is therefore customary. The SARS-CoV-2 diagnosis by RT-qPCR has the following five steps.

1. Saliva sampling has the advantage in that individuals can collect it freely, thus reducing problems of safety management. SARS-CoV-2 viral RNA can be obtained by extracting and purifying using saliva from the subject.
2. Virus inactivation is treated by detergent such as a Tween 20 or a Triton X- 100, or by heating it from 65 to 95°C for several minutes.
3. Extraction and purification for viral RNA in saliva is processed using commercially available extraction and purification kit.
4. RT- qPCR of viral RNA was processed by one-step RT- qPCR.
5. The result is judged from the *Ct* value of the amplification curve of RT- qPCR.

If the cutoff value is 40 cycles or more, it is judged as negative.

### Features of New RT-qPCR Method

The challenges of PCR testing for SARS-CoV-2 infections involve ease of operation, speed with simultaneous processing of multiple samples, accuracy in determining false positives and false negatives, and ensuring the safety of the test result information. About 90% or more of the volume of the PVA sponge is pores, and it is very hydrophilic material. In the medical world, examples of application of culture of bone cells and hepatocytes have been reported^20^. In our case, the PVA sponge was used in order to easily detect the virus in saliva. We have a patent for our technology using a small piece of PVA sponge treated with a biological specimen such as blood or saliva to directly apply PCR master mix solution^21^. The SARS-CoV-2 PCR test kit manufactured by FUJIFILM Wako Pure Chemical has included the primer and probe sets for detecting SARS-CoV-2 viral RNA and the internal control human-derived housekeeping gene *RPL13A*. Generally, the *RPL13A* mRNA must be always detected in saliva, accordingly, the accuracy of false positive or false negative judgment of viral RNA measurement results can be improved. To reduce cost and extend reagent usage, we performed RT-qPCR reactions at half the suggested reaction mix volume.

### Development of a Direct Saliva-to-RT-qPCR Process for Detection of SARS-CoV-2

In general, saliva is comprised of constituents that may hinder virus detection by RT-qPCR, such as degradative enzymes. We sought to identify the conditions under which we could take advantage of the many positive factors of saliva that could be utilized while overcoming the potential limit of detection on this collection medium. For the optimization phase of this study, we utilized processes for inactivated SARS-CoV-2 by a heating method (70°C 15 min).

Finally, to confirm consistency, 25 clinical specimens collected from individuals with suspected COVID-19 symptoms at Showa University were mutually verified by a different preprocessing method using SARS-CoV-2 Detection Kit -Multi- (TOYOBO Co., Ltd., Japan).

### Linear Regression and Limit of Detection for SARS-CoV-2 Viral RNA

The linearity of the *Ct* values in DW and heating saliva is shown in Fig. 3. The concentration gradients of the SARS-CoV-2 viral RNA STDs ranged from 2.7×10^5^ copies/μL to 2.7×10^2^ copies/μL. The coefficients of determination (*R^2^*) of the standard curves for the DW was 0.992 (Fig. 3B). As shown in Fig. 3D, the standard curves *R^2^* of the saliva preparations for heat inactivation was 0.991.

**Fig. 3.**
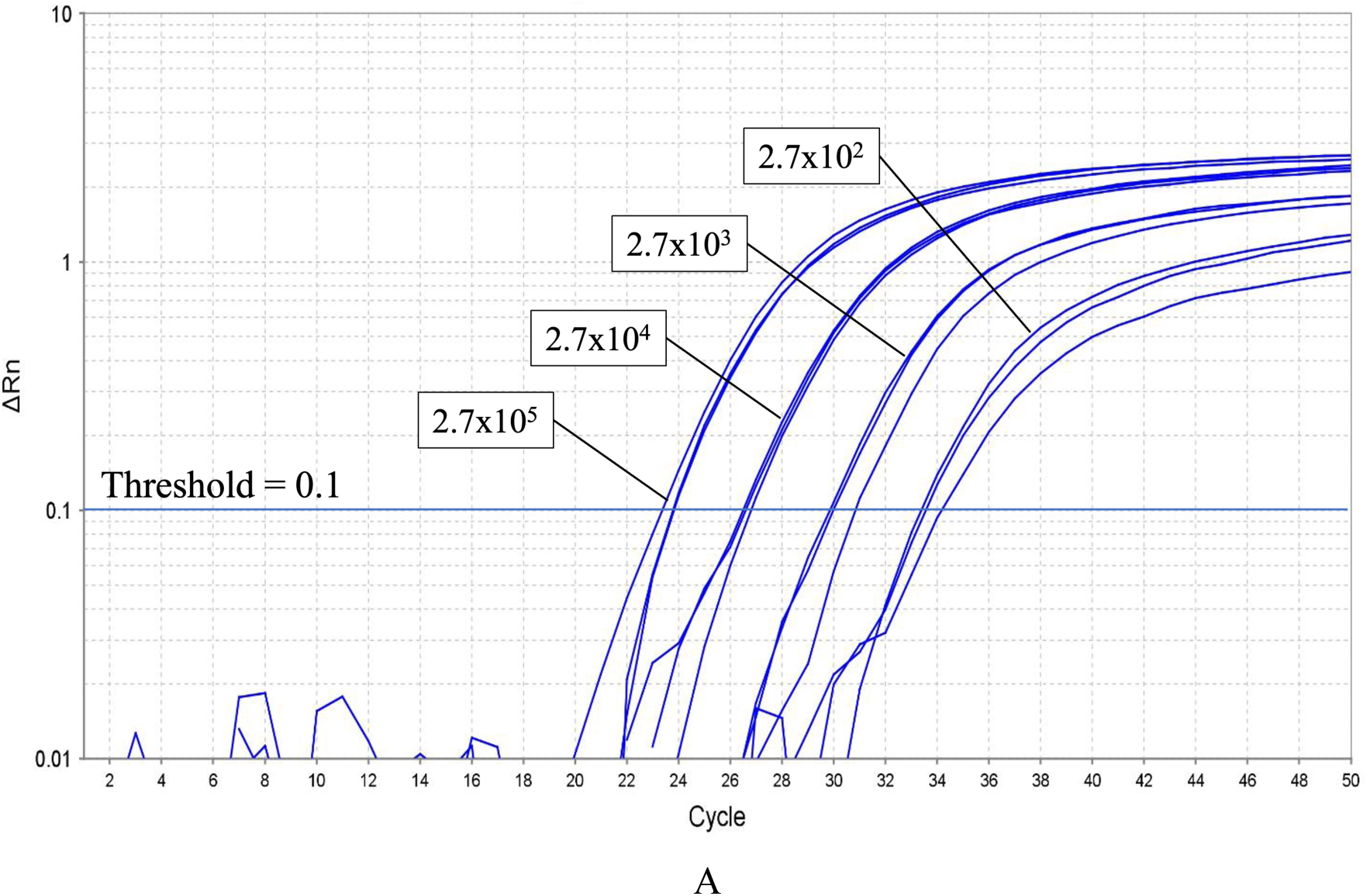
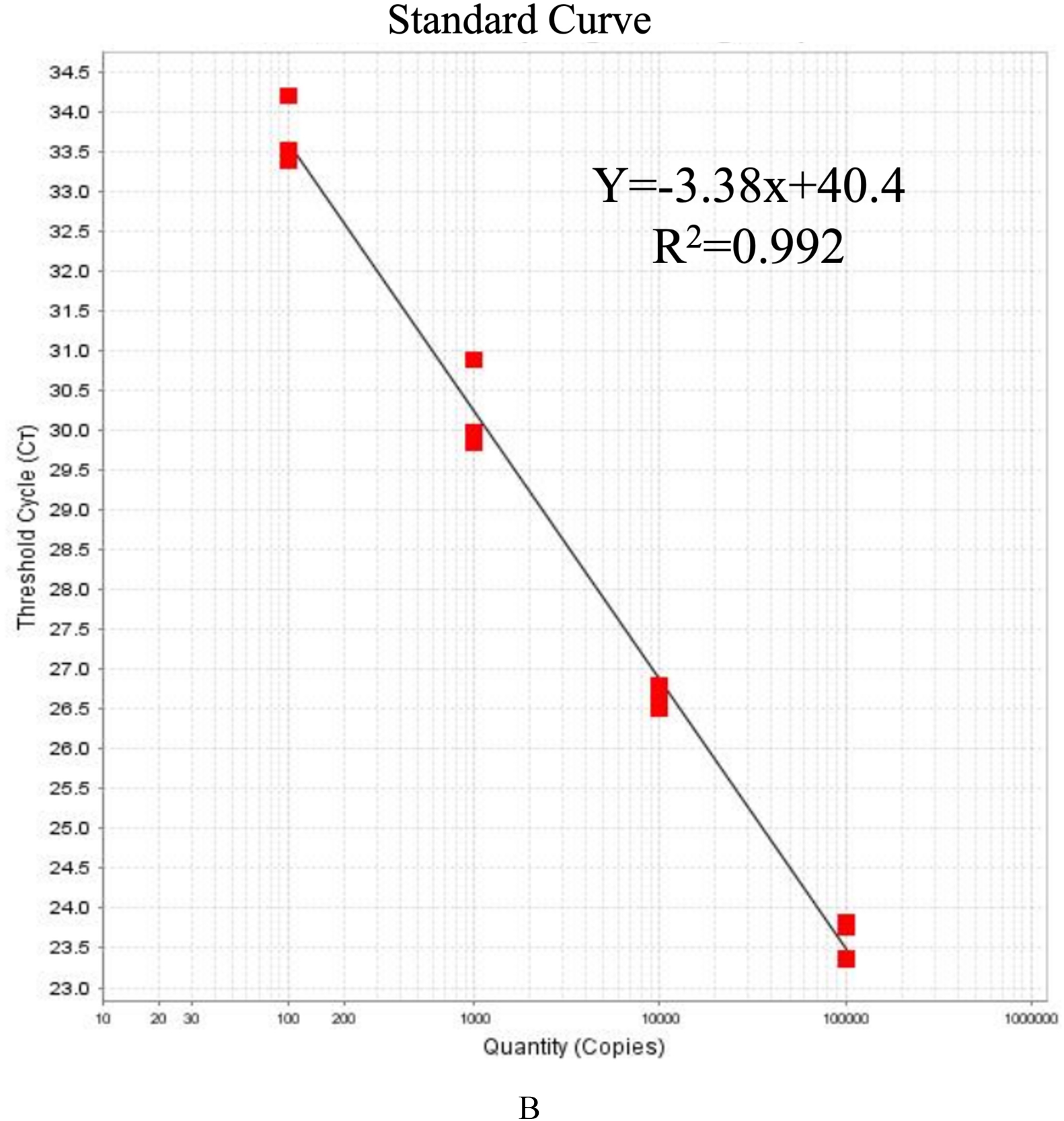
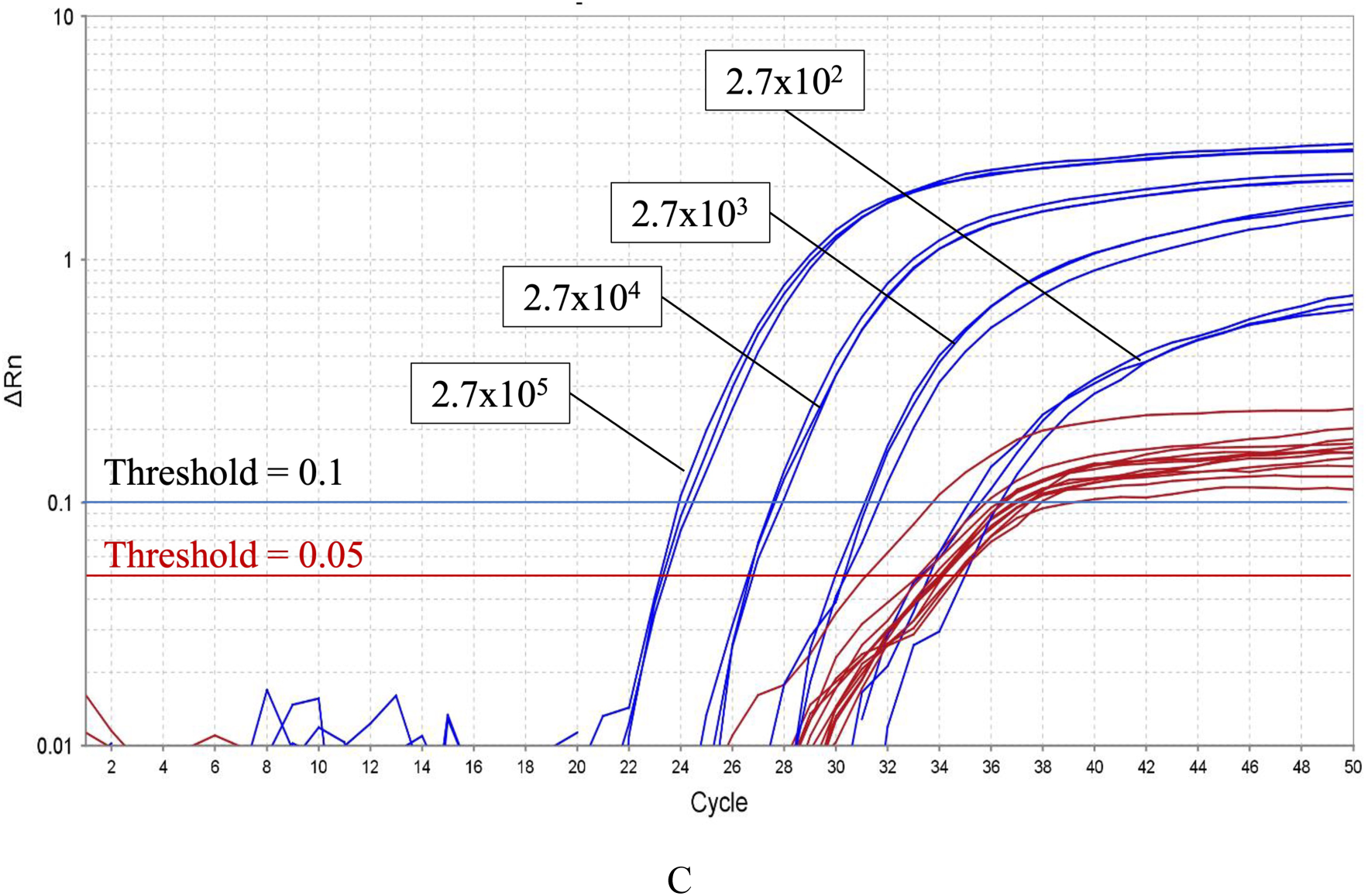
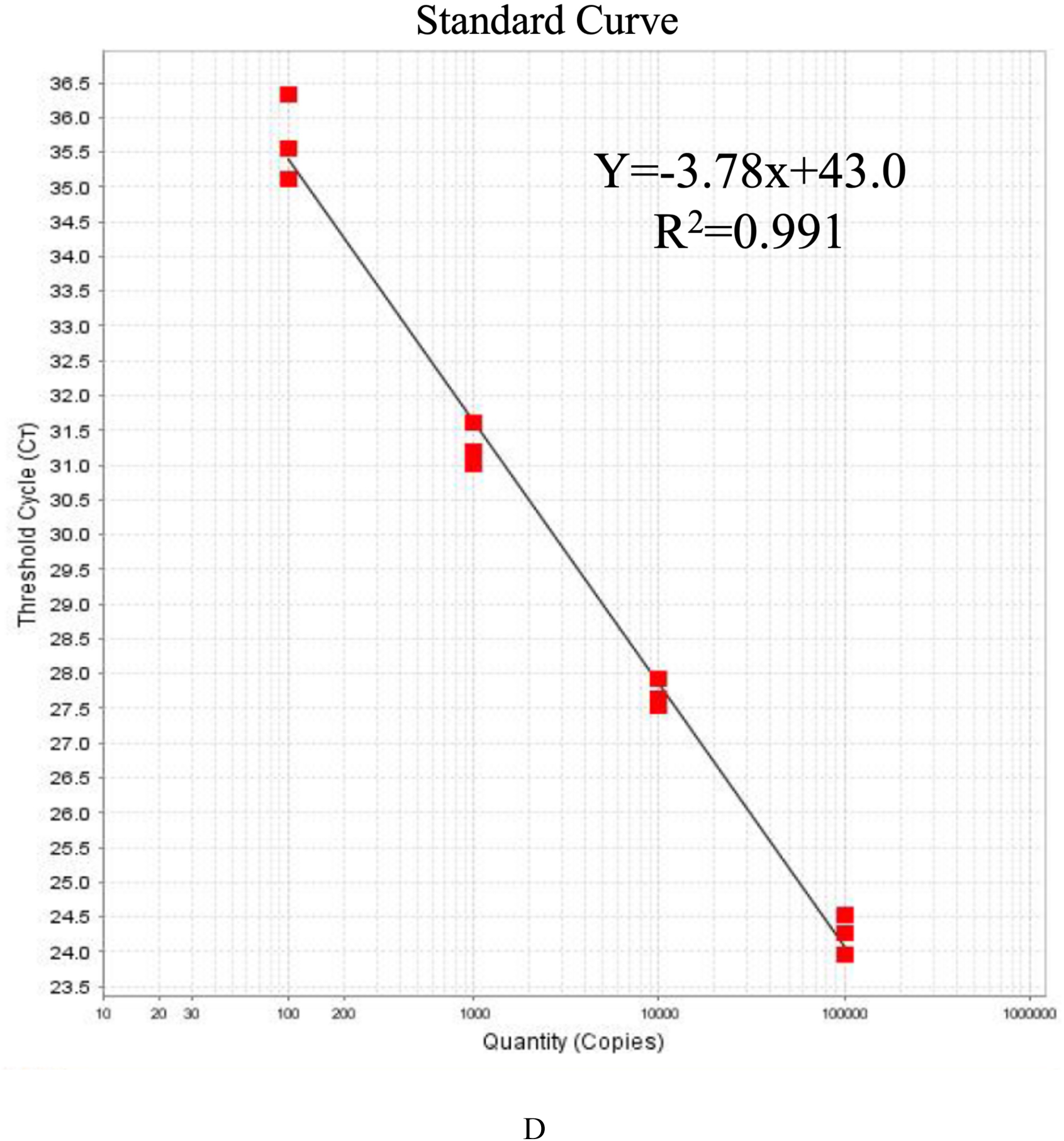
Linear Regression Direct RT-qPCR Analysis for SARS-CoV-2 viral RNA in Distilled Water (DW) and Heating Saliva. A: Amplification Curve in DW. B: Standard curves in DW. C: Amplification Curve in Heating saliva. D: Standard curves in Heating saliva. X axis indicates concentration of SARS-CoV-2 viral RNA STDs (2.7×10^5^, 2.7×10^4^, 2.7×10^3^ and 2.7×10^2^ copies/μL), and Y axis indicates the quantification cycles (*Ct* value) performed by the QuantStudio™ 12K flex real-time PCR system TaqMan^®^ Genotyper^TM^ software.

We assumed that standard curve construction with DW could be useful in the calibration of viral concentrations in saliva samples and in estimating their infectivity. The evaluation of linearity results possessing excellent quantitative ability of N1 and N2 gene analysis by direct RT-qPCR with PVA sponge samples up to 10^2^ copies/μL or less at SARS-CoV-2 viral RNA concentration, which is clearly shown in Fig. 3. The average *Ct* value of 2.7×10^2^ copies/μL-DW samples on PVA sponge was 33.7 cycles ± 0.4. The limit concentration of SARS-CoV-2 viral RNA STDs copy number in DW was supposed to be less than 10 copies/μL by a calibration curve (Fig. 3A and B) as a cut-off *Ct* value of 40 cycles. Furthermore, sensitive detection was demonstrated from the direct RT-qPCR analysis results using the PVA sponge. Accordingly, the *Ct* value of the amplification curve SARS-CoV-2 viral RNA at each concentration was approximately equivalent. Moreover, this study suggests that direct PCR analysis using a PVA sponge was potentially useful for quantitative analysis of SARS-CoV-2, even at very low concentration levels in human saliva as shown in Fig. 3C and D. Also, the *Ct* value of the internal control human-derived housekeeping gene *RPL13A* is shown between 32 and 35 cycles.

### Stability Test in Saliva Containing SARS-CoV-2 Viral RNA on the PVA Sponge

This study shows the stability of the heat treatment method for the quantitative analysis of SARS-CoV-2 viral RNA using human saliva samples. Sample preparations using heating saliva on the PVA sponge and the *Ct* value of the SARS-CoV-2 viral RNA STDs show the higher than that of DW. Even under the sample preparations at 4°C, we thought that the inactivated SARS-CoV-2 RNAs had been decomposed by RNase enzymes in the heating saliva. From these experiments, we have confirmed that SARS-CoV-2 viral RNA on the PVA sponge is stable. In heating saliva samples, when the liquid specimens after 24 h (Day 1) were directly RT-qPCR analyzed, the effect of RNase enzymes in saliva was observed in the *Ct* value as shown in Table 1 compared with that of the PVA sponge. Accordingly, after inactivation, the direct RT-qPCR analysis using liquid biopsy should be analyzed as soon as possible. In the case of direct RT-qPCR analysis using inactivated saliva, strict sample management is required.

**Table 1.** Stability Test of SARS-CoV-2 Viral RNA STDs using the PVA Sponges vs. Liquid Specimens.

| Copy No.<br>copies/μL | DW (*Ct*) | | | Heating Saliva (*Ct*) | | | |
| --- | --- | --- | --- | --- | --- | --- | --- |
|  | Liquid |  | PVA | Liquid |  | PVA |  |
|  | Day 0 | Day 1 | Day 1 | Day 0 | Day 1 | Day 1 | Day 6 |
| $2.7 \times 10^5$ | 23.2±0.1 | 24.5±0.4 | 23.6±0.2 | 24.3±0.4 | 28.1±0.2 | 24.3±0.3 | 24.1±0.5 |
| $2.7 \times 10^4$ | 27.4±0.1 | 27.8±0.8 | 26.6±0.2 | 28.1±0.3 | 32.1±0.5 | 27.7±0.2 | 27.2±0.4 |
| $2.7 \times 10^3$ | 30.6±0.3 | 32.8±0.3 | 30.2±0.6 | 31.5±0.1 | 43.1±1.3 | 31.3±0.3 | 31.6±0.6 |
| $2.7 \times 10^2$ | 35.0±0.3 | 36.2±0.4 | 33.7±0.4 | 38.5±1.1 | ND | 35.7±0.6 | 35.8±0.9 |

The stability of the test of the SARS-CoV-2 viral RNA STDs on a PVA sponge using heating saliva, whose preparation was discussed in the previous section, has been observed by direct RT-qPCR for 6 days at room temperature. As shown in Table 1, the SARS-CoV-2 viral RNA STDs on the PVA sponges were very stable, because the *Ct* values for Day 1 were almost the same as those of Day 6. Accordingly, we thought that SARS-CoV-2 viral RNA STDs prepared using PVA sponge could be used as a positive control for PCR testing of COVID-19 for detecting SARS-CoV-2 viral RNA and the internal control human-derived housekeeping gene *RPL13A*.

### Unique Sample Pooling Method Using PVA Sponge

Given the pandemic, the challenges of RT-qPCR assay for SARS-CoV-2 are mainly concentrated in the operation, which is to timely process multiple samples and to judge false-positive/false-negative accurately. By collecting saliva using a PVA sponge, we have overcome these operational issues, and also this method can skip conventional sample preparation and purification procedures. A standard sample pooling method can be time consuming, costly, and the expected positive rate of the sample can be low^4, 5^. Compared to traditional methods, the decrease in sensitivity due to dilution when the solution is added can be prevented by testing using pieces of dried PVA sponges, and this method does not cause any optical path obstruction as shown in Fig. 4 and Table 2.

**Fig. 4.**
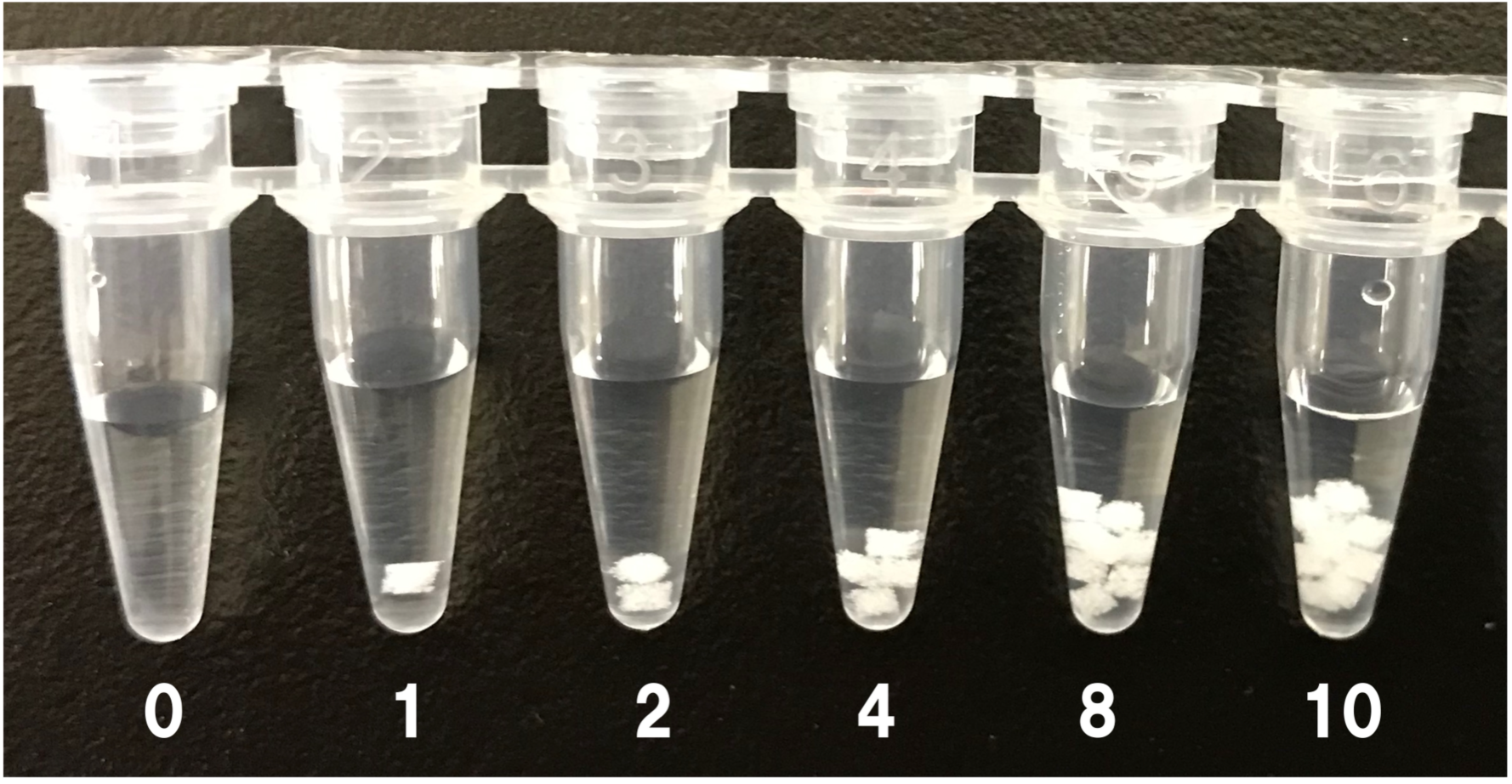
Number of PVA Sponge Disks in Master Mix Solution.

**Fig. 5.**
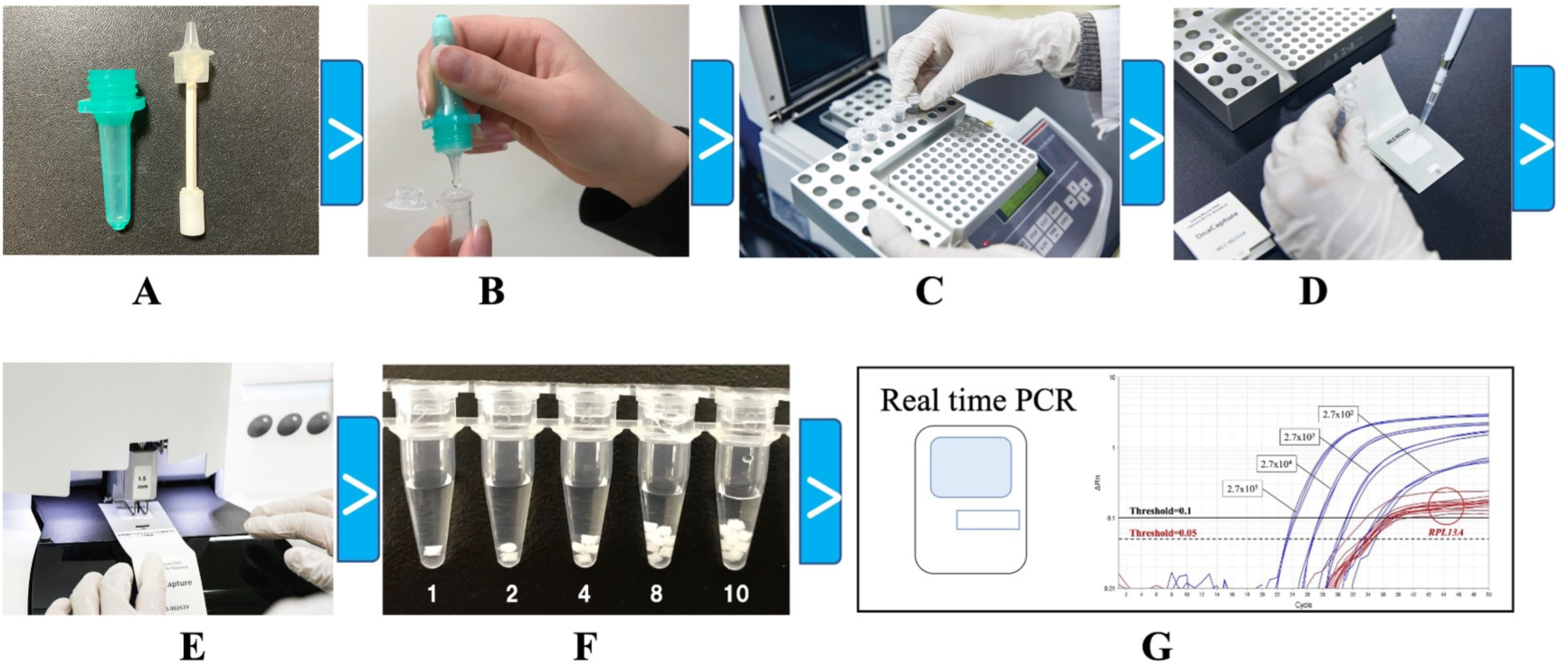
Process from Saliva Sampling and Preparation to Direct RT-PCR A: The subject holds in their mouth until the sponge sufficiently soaked with saliva. B: Transfer saliva by squeezing into a tube. C: Heat inactivation for 15 minutes at 70°C. D: Inactivated saliva soaked on to the PVA sponge. E: After drying, punch out the PVA sponge. F: The number of PVA sponge pieces in a 40µL RT-qPCR master mix solution. G: Direct RT-qPCR by real time PCR system.

**Table 2.** The Change of *Ct* Value using the Sample Pooling Method.

| No. of Positive Control |  | Ten samples Pooling in a Tube |  |  |
| --- | --- | --- | --- | --- |
| A | *Ct* value | A | B | *Ct* value |
| 1 | 31.3 | 1 | 9 | 31.1 |
| 2 | 29.9 | 2 | 8 | 30.6 |
| 4 | 29.7 | 4 | 6 | 29.3 |
| 8 | 28.7 | 8 | 2 | 28.1 |

2 A: No. of Positive Standard PVA sponge disks (1.0 mmΦ as a positive control. The PVA 3 sponge specimens, in which 100 μL of standard SARS-CoV-2 DW solution $2.7 \times 10^3$ 4 copies/μL was added dropwise to the PVA sponge on a UBS card and dried overnight at 5 room temperature, were added directly into the PCR tubes.
6 B: No. of Blank PVA Sponge disks (1.0 mmΦ) as a negative control. *Ct* value: the 7 quantification cycles.

The *Ct* value changes with the number of PVA sponge discs, in which these changes in the *Ct* value are observed in accordance with PCR principles. Furthermore, in our case, since the *Ct* value did not change even when the negative control disk was added, no optical path inhibition was observed in this sample pooling method using a PVA sponge by direct RT-qPCR. Using this method, if ten samples are pooled in a 96-well PCR plate, as many as 960 samples can be tested at the same time. Therefore, when measuring many samples with a low positive rate, it would be possible to improve the efficiency of the test, lessen the time required, and the cost of performing PCR assays.

### General Saliva Sampling and Preparation Using PVA Sponge

Fig. 5 shows an overview of PCR testing at the Mukogawa Women’s University PCR Center (Nishinomiya, Hyogo, Japan). Since its opening in April 2021, more than 8,000 saliva samples regarding SARS-CoV-2 infection have been measured at the center over the past year, and PCR testing has been instrumental in the progress of classes and extracurricular activities on campus and in maintaining the health of elderly residents in nearby facilities.

This unique direct RT-qPCR based method has been performed as follows. First, the subject receives a sampling kit, which includes the Trust Catcher (Trust Medical Co. Ltd. Japan) and the Eppendorf tube. The Trust Catcher is maintained in the subject’s mouth for a few minutes until the sponge is sufficiently soaked with saliva. The saliva is then transferred to the Eppendorf tube after the sponge rod is pushed into the container. The saliva in the Eppendorf tube is heat-treated in a heat block at 70 °C for 15 minutes. The 100 µL of the inactivated saliva samples were dropped on the PVA sponge and then dried in the class II biological safety cabinet at room temperature for at least 3 h with silica gel. These operations of heating of the saliva and drying on the PVA sponge completely lose infectivity of the COVID-19. Small pieces of the specimens (1.5 mm<!) were punched out from the PVA sponge using the Wallac DBS Puncher (PerkinElmer Co., Ltd., USA), then automatically dropped directly into the PCR tubes. Each tube was filled with RT-qPCR master mix solutions, which were adjusted according to the instruction manual. RT-qPCR tests were performed using the QuantStudio™ 12K Flex Real-Time PCR System (Thermo Fisher Scientific Co., Ltd.). The assays were performed with the sample pooling method as mentioned above^22^.

## Conclusion

This report demonstrated that our direct RT-qPCR analysis method using a PVA sponge disk enables the quantitative, real-time PCR analyses of SARS-CoV-2 concentrations in saliva samples without any RNA extraction and purification. This sensitive quantitative detection can contribute to restricting the spread of SARS-CoV-2 infection because it can detect low viral numbers, i.e., less than 10 copies/μL, in saliva samples obtained from early-stage COVID-19 patients or asymptomatic individuals.

The PVA sponge is a material with hydrophilic and hydrophobic properties, and it can be presumed that it has a high affinity for enzymes. We have not examined the advantage of using the PVA sponge in detail, but we consider that the action of RNA degrading enzyme eg RNase is suppressed by its interaction with the PVA sponge, and the reaction progress of viral RNA degradation is hard to occur even in samples like saliva.

We have developed a novel RT-qPCR testing procedure by using a PVA sponge without purification, which can be utilized for the detection of SARS-CoV-2. Finally, 25 subjects (8 positive and 17 negative) allowed us to confirm that results were consistent with our suggested method (data not shown). Therefore, it is our hope that this basic study described herein encourages further validation of our method by clinical researchers who may be interested in this simple, reliable, and cost-effective method.

In summary, described herein is a sensitive diagnostic method for SARS-CoV-2 that is operationally simple, evaluates a clinically relevant infectious biological specimen, is appropriate for large scale repeat testing, is cost-effective, and can be readily adopted by a BSL-2 laboratory. Large scale SARS-CoV-2 testing will be a powerful weapon in preventing the spread of this virus and helping to control the COVID-19 pandemic.

## Ethics statement

This study did not require IRB approval because only non-identifiable self-collected specimens were used.

## Author contributions

K.K. conceived the study, conducted experiments, analyzed the data, and wrote the manuscript.

## Data Availability

All data produced in the present study are contained within the manuscript. Additional raw data are available from the corresponding author upon reasonable request.

## Acknowledgments

We would like to thank Professor Dr. Yuji Kiuchi et al. at Showa University for the providing clinical specimens, and Ms. Youtaro Yamamoto et al. at FUJIFILM Wako Pure Chemical Co., Ltd. for the PCR technical support. This study was performed with the support of university-owned testing equipment utilization promotion project for the Ministry of Education, Culture, Sports, Science and Technology, Japan (MEXT).

## Author contributions

K.K. contributed in evaluating, preparing and interpreting the data, designed, managed and supervised the project. All authors edited the manuscript to its final form. M.K. performed RT-q PCR experiments. Both T.S. performed preparation of standard and organization for inactivation of SARS-CoV-2. K.S and A.I. conceived and supervised the project

